# Pan-cancer analysis reveals unique molecular patterns associated with age

**DOI:** 10.1101/2020.08.30.20184762

**Authors:** Yajas Shah, Akanksha Verma, Andrew Marderstein, Bhavneet Bhinder, Olivier Elemento

**Affiliations:** Caryl and Israel Englander Institute for Precision Medicine, Weill Cornell Medicine, New York, NY 10021, USA; Institute of Computational Biomedicine, Weill Cornell Medicine, New York, NY 10021, USA; Physiology, Biophysics and Systems Biology Graduate Program, Weill Cornell Medicine, New York, NY 10065, USA; Tri-Institutional Program in Computational Biology and Medicine, Weill Cornell Medicine, New York, NY 10065, USA

## Abstract

Older age is a strong risk factor for several diseases, including cancer. In cancer, older age is also frequently associated with a more aggressive, treatment-refractory tumor phenotype. The etiology and biology of age-associated differences among cancers are poorly understood. To address this knowledge gap, we sought to delineate the differences in tumor molecular characteristics between younger and older patients across a variety of tumor types. We found that tumors in younger and older patients exhibit widespread molecular differences. First, we observed that tumors in younger individuals, unlike those in older ones, exhibit an accelerated molecular aging phenotype associated with some hallmarks of premature senescence. Second, we found that tumors from younger individuals are enriched for driver gene mutations resulting in homologous recombination defects. Third, we observed a trend towards a decrease in immune infiltration and function in older patients and found that, immunologically, young tumor tissue resembles aged healthy tissue. Taken together, we find that tumors from young individuals possess unique characteristics compared to tumors in older individuals, which can potentially be leveraged for differential therapeutic strategies.

## Introduction

Aging refers to the decline in physiological functions with time, resulting in an increased risk of disease. Although advances in healthcare have extended lifespan substantially, cancer continues to be a significant contributor to global mortality. The United States projects 1.8 million new cancer cases and 600,000 cancer-related deaths in 2020(Siegel et al., 2020). Several diseases, including cancer, are typically diagnosed in older populations. Older patients frequently have worse outcomes. The molecular correlates of such age-associated differences are not known.

Moreover, the incidence of cancer in young adults is increasing at an alarming rate (Ahnen et al., 2014; Anders et al., 2009; Ben-Aharon et al., 2019). In the United States, one in twenty-nine males and one in seventeen females under the age of 49 are likely to develop cancer as per a recent report(Siegel et al., 2020). Tumors in the breast, colon, rectum, genital tract, skin, connective tissue, and thyroid gland are the most common in this age group (Bleyer et al., 2008; Jemal et al., 2010; Siegel et al., 2020).

It is unclear whether tumors in younger and older adults have distinct biology. Breast cancers in young adults are typically larger, often harbor the triple-negative phenotype, and are associated with mutations in *BRCA1, BRCA2*, and *TP53* (Anders et al., 2009; Lalloo et al., 2006). Colorectal cancers in young adults may have a high degree of microsatellite instability (MSI) and are enriched for mutations in *MYCBP2, BRCA2, PHLPP1, TOPORS*, and *ATR* compared to older patients(Tricoli et al., 2018). While several lines of evidence suggest that cancers in young adults show unique histology and survival heterogeneity, their biology has not been well characterized (Bleyer et al., 2008; Keegan et al., 2016).

In this study, we conduct an unbiased analysis of primary tumors in The Cancer Genome Atlas (TCGA) to understand the biology of cancers in younger vs. older individuals. Furthermore, we sought to elucidate genomic, epigenomic, and transcriptomic aberrations in younger and older patients and contrast them with healthy tissue. Understanding the unique biology of tumors in younger and older patients may lead to personalized therapeutic strategies and the development of additional biomarkers.

## Results

### Identification of age-associated cancers

We first sought to identify cancers with significant age-dependent outcomes in TCGA, reasoning that the biology underlying the difference in outcomes may be more acute and interpretable in such cancers compared to cancers with no age-associated outcome differences. These cancer types were identified by a two-step filtration process involving a Cox-proportional hazards model as well as differential gene expression of primary tumors. First, tumor types in which the overall survival (OS) of a patient stratified by age at diagnosis were selected by Cox regression using age at diagnosis as a covariate. This identified thyroid carcinoma (THCA), low-grade gliomas (LGG), acute myeloid leukemia (LAML), uterine corpus endometrial carcinoma (UCEC), glioblastoma multiforme (GBM), breast invasive carcinoma (BRCA), bladder urothelial carcinoma (BLCA), colon adenocarcinoma (COAD), kidney renal cell carcinoma (KIRC), skin cutaneous melanoma (SKCM), ovarian carcinoma (OV) and head and neck squamous cell carcinoma (HNSC) as tumor types in which overall survival was inversely related to age (FDR < 0.01, **Fig. 1A**, Supplementary Table 1). We divided samples from each of these tumor types into quartiles based on their age of diagnosis. The first and fourth quartile served as younger and older age groups, respectively (**Supplementary Fig. 1A**, Supplementary Table 2). Next, we identified tumors with an age-dependent molecular phenotype by differential gene expression (DGE) of younger and older groups. Tumor types that had greater than 1% of genes differentially expressed between the age groups were deemed to show an age-associated molecular phenotype (FDR < 0.05, **Fig. 1B**) and were selected for all further analyses. Tumor types in this group included BRCA, LGG, UCEC, OV, THCA, and COAD.

**Figure 1:**
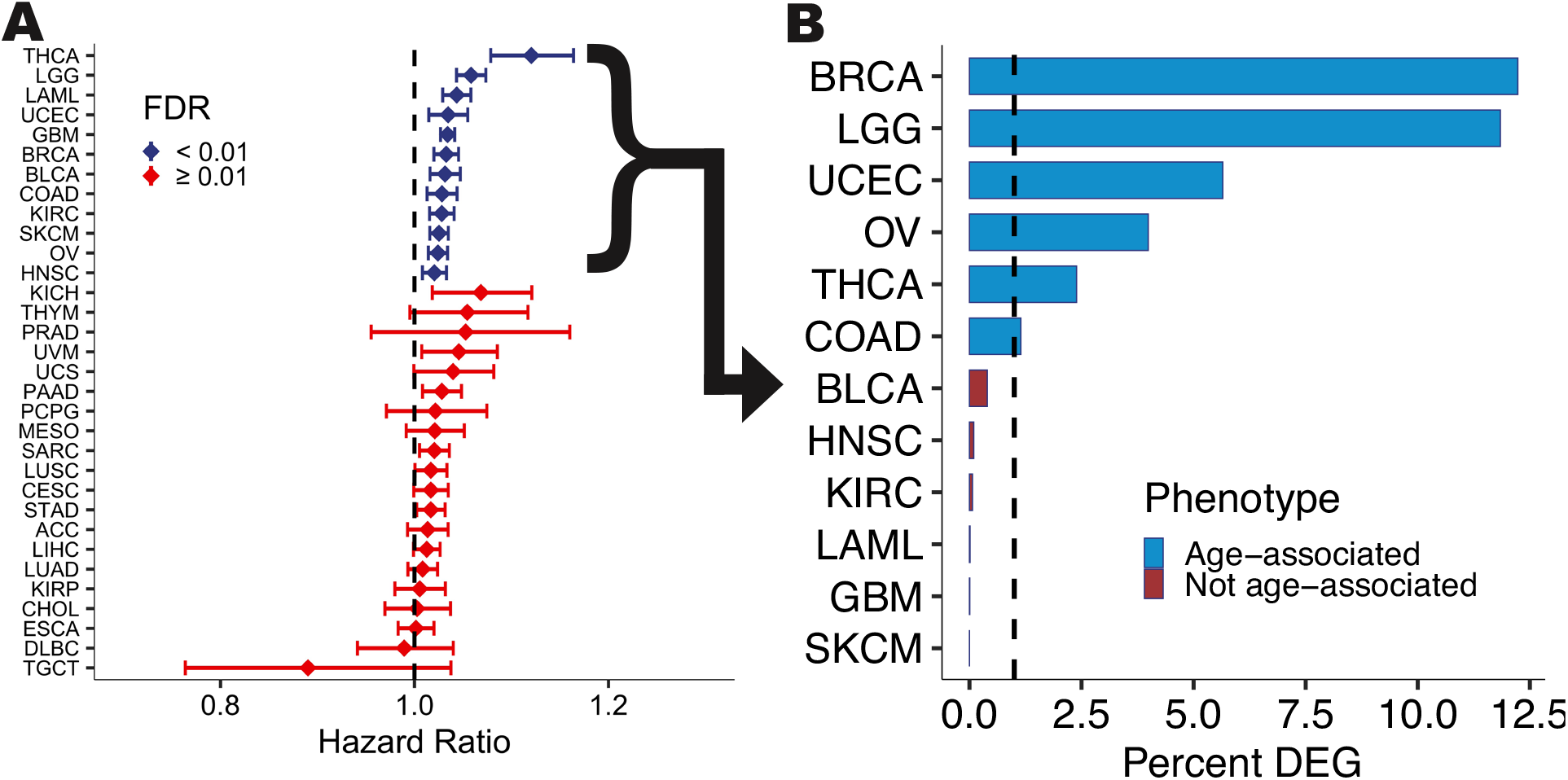
Tumor type selection. (A) Hazard ratios (HRs) for overall survival as a function of tumor diagnosis age. HRs greater than 1 are associated with decreased survival probability in older patients. Tumor types in dark blue have significantly different outcomes based on age (FDR < 0.01) (B) Differentially expressed genes (DEG) between younger and older patients (FDR < 0.05) from tumor types with HR > 1. The bar plot indicates the fraction of DEG. Tumor types with HR > 1 and DEG > 1% were termed age-associated cancers (light blue).

We wondered whether younger age groups would be associated with early-stage tumors. To our surprise, we did not find any association between age groups and tumor stage in most tumor types of interest; THCA was the only tumor type that displayed this trend. In THCA, Stage I tumors were associated with younger patients, while higher stages were associated with older patients (FDR < 0.05, **Supplementary Fig. 1B**).

### Aging drives proliferation and immune dysfunction in cancer

We sought to explore the association between age and functional aspects of tumor progression. To do this, we correlated gene expression levels of commonly used tumor progression markers (*MYBL2, TOP2A, PLK1, CCND1, PCNA*, and *MKI67*) with patient age. We found that in most tumor types, higher gene expression levels of these markers were inversely correlated with age, that is, more highly expressed in younger patients, indicating that tumors from younger patients may be more aggressive than those in older ones (FDR < 0.05, **Supplementary Fig. 1C**). To further explore this, we sought out to obtain a more detailed understanding of transcriptional changes associated with aging in cancer using differential gene expression. Of the tumor types we analyzed, we found that the BRCA, LGG, and UCEC had the most differentially expressed genes between younger and older patients (**Fig. 1B**). In line with the greater expression of proliferation markers in young patients, we found that gene set enrichment analysis of young vs. old DGE results for KEGG pathways associated with tumor growth linked younger patients to a proliferative phenotype (FDR < 0.05, **Supplementary Fig. 1D**).

Overall, we found that genes differentially expressed with aging show a high degree of overlap across tumor types, suggesting that at least some of the biology underlying aging processes in cancer is consistent across tissue types. Using pairwise Fisher’s exact tests, we found that there is a greater overlap between overexpressed genes in younger patients across tumor types as compared to older ones (FDR < 0.05, **Supplemental Fig. 1E**). Seventeen genes were over-expressed in younger cohorts across more than three tumor types. These genes included *ZNF518B, EDNRA, GMEB1, PPP1R10*, and *FERMT1* and have been linked to tumor growth, metastasis and poor survival in a variety of cancers(An et al., 2019; Gimeno-Valiente et al., 2019; Kavela et al., 2013; Laurberg et al., 2014; Liu et al., 2017). In contrast, we found ten genes to be over-expressed in old patients across at least three out of six tumor types. These include *COQ3, EYA4, FER1L5, HOXB5, SYS1* and *TSNAX* and have been associated with a variety of cellular functions including mitochondrial function, tumorigenesis, memory formation and protein trafficking(Behnia et al., 2004; Cannon et al., 2005; Jonassen and Clarke, 2000; Lee et al., 2018). Altogether these findings indicate that primary tumors in younger patients have increased expression of tumor proliferation, progression, and metastasis genes. We hypothesized that such tumors while having better outcomes than in older patients, are, in fact, more aggressive but perhaps restrained by a more functional immune system.

To explore this hypothesis, we conducted gene set enrichment analyses of differential expression results. This analysis showed that younger patients were enriched for pathways associated with immune response in BRCA, THCA, OV, and UCEC cohorts (FDR < 0.05, **Fig. 2A**). Interestingly, we found that older patients in the LGG and COAD cohorts displayed the opposite pattern and were enriched for immune-associated pathways compared to younger patients, suggesting that aging-associated effects were tumor type specific. It is possible that this enrichment was associated with inflammaging rather than anti-tumor immunity. Inflammaging refers to inflammation commonly associated with aging and is considered to be one of the several evolutionarily conserved pillars of senescence (Franceschi et al., 2018; Montecino-Rodriguez et al., 2013; Nikolich-Žugich, 2018).

**Figure 2:**
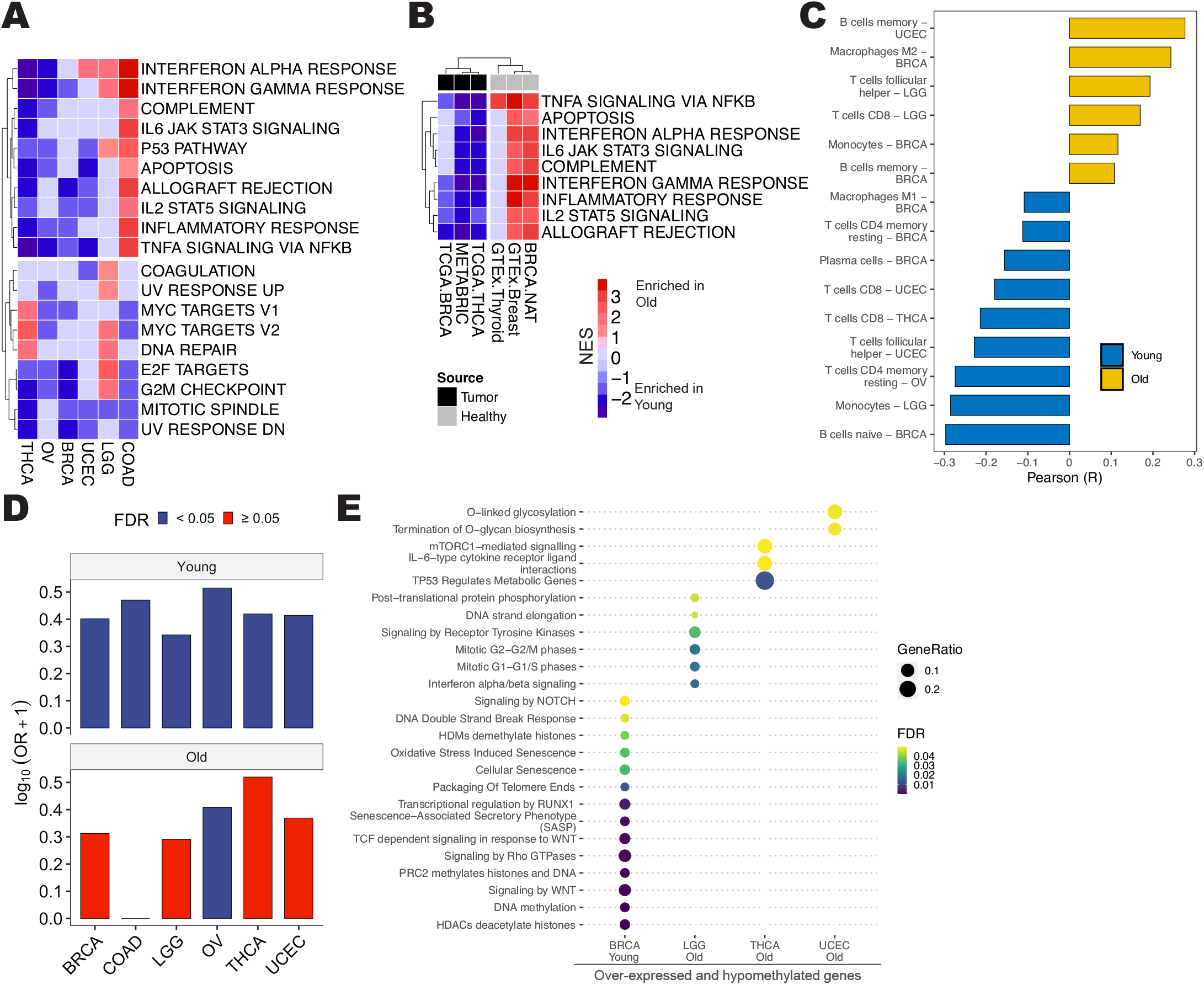
Differential gene expression and methylation links immune function and senescence to young patients. (A) Gene set enrichment analysis of DEG between younger and older patients across tumor types. Pathways in red are associated with older patients, and those in blue are associated with younger patients (FDR < 0.05). (B) Gene set enrichment analysis of DEG between tissue sourced from younger and older cancer patients (black) as well as corresponding healthy individuals (grey). (C) Pearson correlation coefficients of CIBERSORT (relative) scores with age across tumor types (FDR < 0.05). (D) The bar plots show the overlap between hypomethylated and over-expressed genes in young (top) and old patients (bottom). (E) Reactome pathway enrichment analysis of hypomethylated and over-expressed genes. Point size reflects the ratio of the number of genes present in the differential gene list and the number of genes present in the pathway.

Next, we aimed to understand whether the previously mentioned transcriptional changes reflected aging specific to cancer or were simply a consequence of healthy aging. To address this question, we compared young vs. old differential gene expression results from TCGA with the results from an identical analysis performed on healthy tissue (sourced from the Genotype-Tissue Expression project (GTEx) project). This analysis allowed us to identify tissue-type specific genes associated with aging (**Supplementary Fig. 1F**). *EYA4*, a gene associated with hearing loss and cardiomyopathy(Abe et al., 2018), was over-expressed in older individuals regardless of cancer status in four out of six tissue types. Similarly, *HENMT1, NOA1*, and *ZNF518B* were over-expressed in younger individuals in three out of six tissue types. These genes have been linked to piRNA methylation(Begik et al., 2020), mitochondrial function(Kolanczyk et al., 2011), and tumorigenesis(Gimeno-Valiente et al., 2019).

We included the METABRIC dataset (breast cancer) and further investigated these results at the level of activated pathways using GSEA. This analysis showed that the relationship between aging in cancer and healthy aging was tumor type-dependent (**Fig. 2B**). We found that in breast cancer, immune pathways (allograft rejection, IL-2 signaling, inflammatory response, interferon a/y response, complement, IL-6 signaling, apoptosis, and TNFα signaling) were overall upregulated in older healthy donors and, as shown above, in young cancer patients (FDR < 0.05, **Fig. 2B**). While thyroid cancer showed a similar phenotype of pathway activation, other tumor types did not display this pattern. Thus, patterns of pathway activation suggest that younger breast and thyroid cancer patients resemble immunological phenotypes of aged corresponding healthy tissue, indicating dysregulation of the aging process in cancer (**Fig. 2B**).

To obtain a more granular understanding of immune function, we correlated immune cell infiltration (estimated using CIBERSORT(Newman et al., 2015)) with age tumor diagnosis. We found that the correlation between age and specific cell types varied between tumor types (**Fig. 2C**). While we did not observe any strong correlations involving specific cell populations, we found that immune cells with anti-tumor potential were predominantly associated with younger populations. Indeed we found that CD8^+^ T cells were associated with younger THCA and UCEC patients. Similarly, CD4^+^ resting memory T cells were associated with younger BRCA and OV patients. M1 macrophages and antibody-secreting plasma cells were linked to younger BRCA patients (FDR < 0.05). Interestingly, older BRCA patients showed an increase in M2 macrophages, and LGG patients were associated with CD8^+^ T cells (FDR< 0.05). Memory B cells were associated with older patients in BRCA and UCEC (FDR < 0.05). While the role of B cells in cancer is tumor-type dependent and remains controversial(Garaud et al., 2019; Zhang et al., 2016), M2 macrophages are one of the major immunosuppressive species in the tumor microenvironment(Pyonteck et al., 2013). However, increased T cell infiltration has been unequivocally shown to result in a more robust anti-tumor response and better prognosis in multiple tumor types(Hodi et al., 2010; Le et al., 2015; Topalian et al., 2012). These data are concordant with increased enrichment of immune infiltrating cells in young patients.

Taken together, we observed that several genes associated with progression, metastasis, and poor survival outcomes are associated with primary tumors from younger patients. Furthermore, we show that while immune pathway enrichment with aging occurs in a tumor-type specific manner, younger patients typically may harbor a more robust immune response in the tumor microenvironment. This appears to balance the effect of aggressive gene expression profiles (**Supplementary Fig. 1D**) found in these patients.

### Functional analysis of age-associated DNA methylation marks

Most mammalian cells undergo global loss of DNA methylation marks and other epigenetic modifications with aging, which consequently result in transcriptional imbalances(Sen et al., 2016). We hypothesized that a methylation signature of aging would exist in tumors as well. To test this, we identified tumor type-specific differentially methylated genes for the age groups of interest (see Methods). We found that LGG and BRCA had the highest number of differentially methylated genes (**Supplemental Fig. 1G**). Interestingly, a high percentage of genes were hypermethylated in older patients from the COAD, THCA, and BRCA cohorts (**Supplemental Fig. 1G**).

As expected, we found a robust association between hypomethylated and over-expressed genes (**Fig. 2D**). This association was strongest in genes over-expressed in younger patients, where all tumor types showed significant overlaps with hypomethylated genes (FDR < 0.05, **Fig. 2D**). In contrast, there were fewer overlaps between DNA hypomethylation and gene expression of genes that were over-expressed in older populations, where only OV showed this trend. We created gene lists representing the age-stratified (young or old) tumor-type specific intersection of hypomethylated and over-expressed genes. Next, we conducted Reactome pathway enrichment analysis(Yu and He, 2016) of these gene sets to understand epigenetically driven alterations. We found that gene lists from old LGG, THCA, and UCEC patients were enriched for few pathways (phases in mitosis, mTORC signaling, glycosylation), while the gene list from younger BRCA patients, but not older BRCA patients, was enriched for multiple pathways (Supplementary Table 3). Several pathways associated with the gene list obtained from young breast cancer patients were linked to senescence (senescence associated secretory phenotype, cellular senescence, oxidative-stress induced senescence), epigenomic reprogramming (HDACs deacetylate histones, HDMs demethylate histones, DNA methylation), oncogenic signaling (Wnt signaling), and DNA damage response (DNA double-strand break response) (**Fig. 2E**). These features describe aging imbalance and oncogenic processes, which may, in part, explain the aggressiveness of tumors from younger breast cancer patients.

Taken together, we show that genes that are over-expressed in younger cancer patients may be epigenetically controlled. Furthermore, epigenetically controlled pathways associated with young breast cancer patients, but not other cancers, are enriched for senescence, suggesting dysregulated aging in the tumor.

### Tumors from young patients exhibit accelerated molecular aging and are senescent

To understand the extent of aging-imbalance, we characterized the molecular age of a tumor in terms of the gene expression and DNA methylation (DNAm) profiles. We estimated the DNAm age from BRCA, LGG, UCEC, THCA, and COAD TCGA cohorts (Supplementary Table 4) and found a weak correlation with chronological age (R = 0.12, p = 6.6×10^-8^) in primary tumor samples (**Fig. 3A**). We did not estimate the DNAm age of the OV cohort since there were not enough samples for statistical analysis. Interestingly, the DNAm age of healthy normal adjacent tissue from TCGA (NAT) showed a high correlation with actual patient age (R = 0.79, p < 2.2×10^-16^), suggesting that aberrant epigenetic landscape of the tumor alters its epigenetic age. By comparing the donor age with DNAm age, we found that tumors from young and middle-aged patients displayed an accelerated epigenetic age phenotype when compared to NAT (**Fig. 3B**). However, tumors from older patients did not display this phenotype as prominently, where epigenetic age and actual age were much closer, suggesting that epigenetic age acceleration is observed in younger populations. These results are in line with previous work, which showed that very young breast cancer patients had accelerated epigenetic ages(Oltra et al., 2019). Indeed, consistent with the findings of Horvath, 2013, apart from LGG, most tumor types tested exhibited the trend of reduced age acceleration upon aging (**Fig. 3C**).

**Figure 3:**
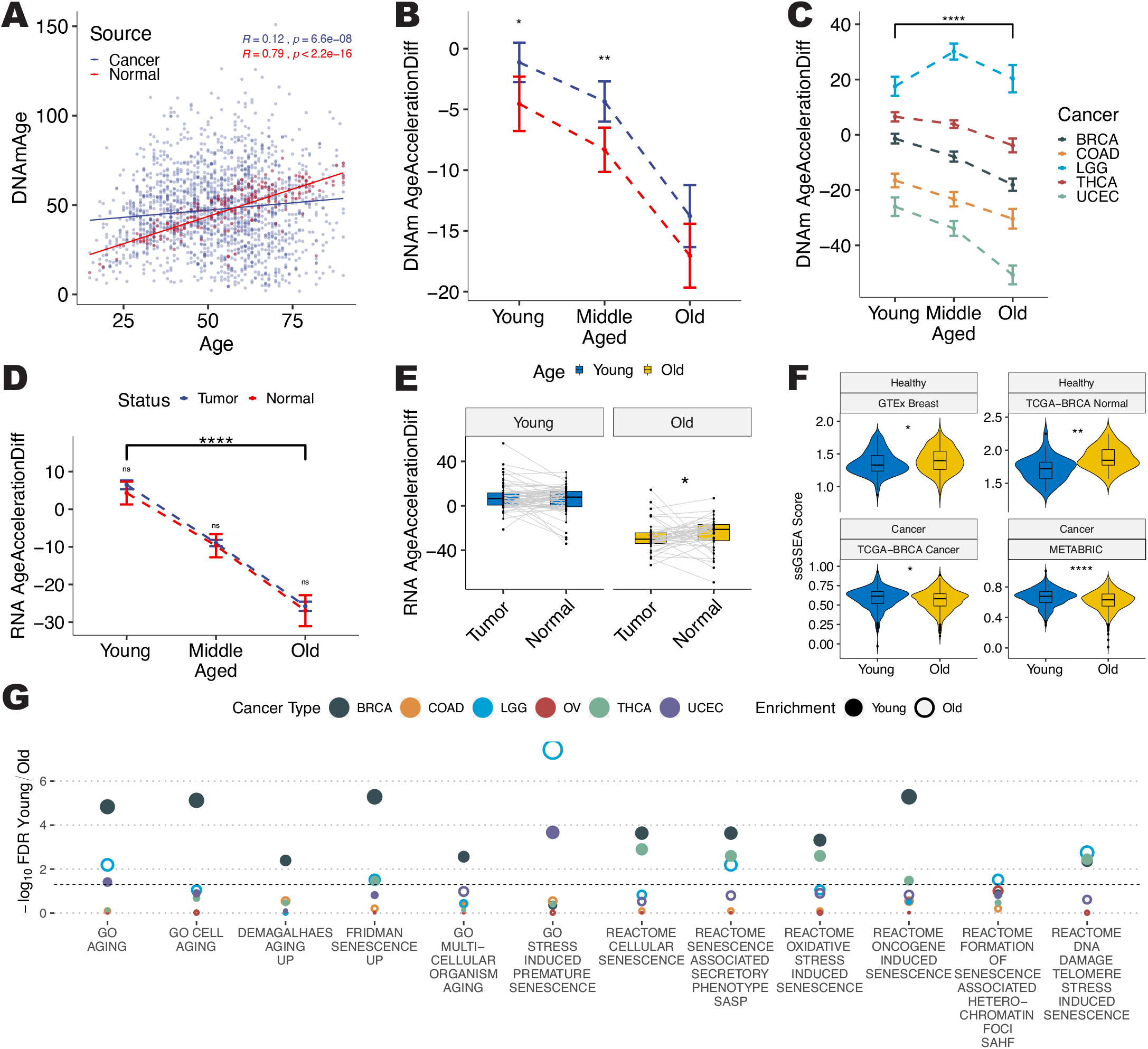
Young patients are associated with senescence and accelerated aging. (A) Scatterplot of DNA methylation age and chronological age in tumor (dark blue) and healthy (red) tissue. (B) The difference between DNA methylation age and chronological age (DNAm age acceleration difference) is plotted against chronological age bins. (C) DNAm age acceleration difference is plotted against age bins for breast cancer (dark green), colon cancer (light brown), low grade glioma (blue), thyroid cancer (dark brown), and uterine endometrial carcinoma (green). (D) The difference between RNA age and chronological age (RNA age acceleration difference) is plotted against age bins for tumor (dark blue) and normal (red) samples. (E) Paired analysis of RNA age acceleration difference in younger and older patients. (F) Single sample gene set enrichment analysis of genes associated with aging in healthy (top) and tumor (bottom) breast samples. (G) The scatter plot shows BH-adjusted p values for differential pathway enrichment analysis between young (filled circles) and old (hollow circles) patients from the BRCA (dark green), COAD (light brown), LGG (blue), OV (dark brown), THCA (green) and UCEC (purple) TCGA cohorts.

After observing this phenotype in DNA methylation, we aimed to recapitulate these findings in gene expression data. In order to do so, we used the framework set up by Ren and Kuan, 2020. Similar to the DNAm age calculator, Ren & Kuan have trained tissue-type specific age predictors for RNA-Seq data. We imputed RNA age using these tissue-specific models for BRCA, COAD, THCA, and UCEC (Supplementary Table 5). The analysis was limited to these tumor types because Ren & Kuan do not provide regression coefficients for other tissue types, or because there were no healthy samples in our dataset. Similar to the DNAm age calculator, the RNA age for healthy tissue correlated with chronological age much better than tumor tissue (**Supplementary Fig. 2**). Interestingly, we observed a similar trend of reduced age acceleration upon aging (p < 0.05, **Fig. 3D**). While we did not observe accelerated aging in tumors from young patients (potentially due to sample size), tumors from old patients had reduced age acceleration when compared with matched healthy tissue (p < 0.05, **Fig. 3E**).

Next, we sought to further examine accelerated aging at the transcriptional level in breast cancer using gene signatures of aging. We focused this analysis on breast cancer since four unique datasets (TCGA-BRCA tumor, TCGA-BRCA normal tissue, METABRIC, and GTEx breast tissue) were available to us. We calculated the transcriptional age of all samples using single sample gene set enrichment analysis (ssGSEA) enrichment of genes upregulated with aging in primary human fibroblasts (mSigDB M8910). We found that younger breast cancer patients have an older tumor phenotype compared to chronologically older patients in the TCGA-BRCA and METABRIC datasets (p < 0.05, **Fig. 3F**). In contrast, healthy breast tissue samples obtained from GTEx and TCGA do not display a transcriptional age acceleration phenotype.

In order to assess these changes in other tumor types, we used differential pathway enrichment signals from an ssGSEA-based analysis. To do so, we estimated ssGSEA enrichment scores for younger and older patients across tumor types independently and subsequently calculated differential enrichment signals using a t-test. Similar to immune pathway activation (**Fig. 2A**), we found that pathways associated with aging and senescence show tumor-type specific patterns (**Fig. 3G**). Interestingly, younger BRCA patients were associated with almost all senescence and aging pathways we could find in MSigDB. Additionally, younger THCA patients were associated with cellular senescence, senescence associated secretory phenotype, oncogene-induced, oxidative stress-induced, and telomere stress-induced senescence along with several aging-related pathways (FDR < 0.05). Similarly, younger OV patients had a greater enrichment score for stress induced premature senescence than old ones (FDR < 0.05). In contrast, LGG was the only tumor type in which these pathways were differentially associated with older patients.

Taken together, we show that molecular age acceleration and senescence are associated with younger patients, rather than older. Since some senescent phenotypes have been shown to promote tumor growth (Fane and Weeraratna, 2020), we show that aggressive tumor phenotypes may be explained, in part, by defunct cellular pathways controlling senescence.

### Age-associated mutational profiles

The majority of human cancers are caused by the sequential alteration of several genes over the course of multiple years(Vogelstein et al., 2013). We compared somatic mutations between tumors from younger and older patients in order to understand age-associated mutational patterns. The most commonly mutated genes in cancers with age-dependent outcomes were *TP53* (39%), *PIK3CA* (23%), *APC* (13%), and *PTEN* (13%) (**Fig. 4A**). While all variants in a cancer driver genes do not have an equal impact in tumorigenesis, there is an increased probability of tumor growth when driver genes carry a larger number of variants(Carter et al., 2009; Torkamani and Schork, 2008). Interestingly, younger patients were enriched for mutations in driver genes *(TP53, ATRX, KMT2C, ARID1A)* with more than one variant (FDR < 0.05, Fisher’s Exact Test). In line with previous research(Chalmers et al., 2017), we found that older patients had a higher tumor mutation burden (TMB) in most age-associated cancers (FDR < 0.05, **Fig. 4B**).

**Figure 4:**
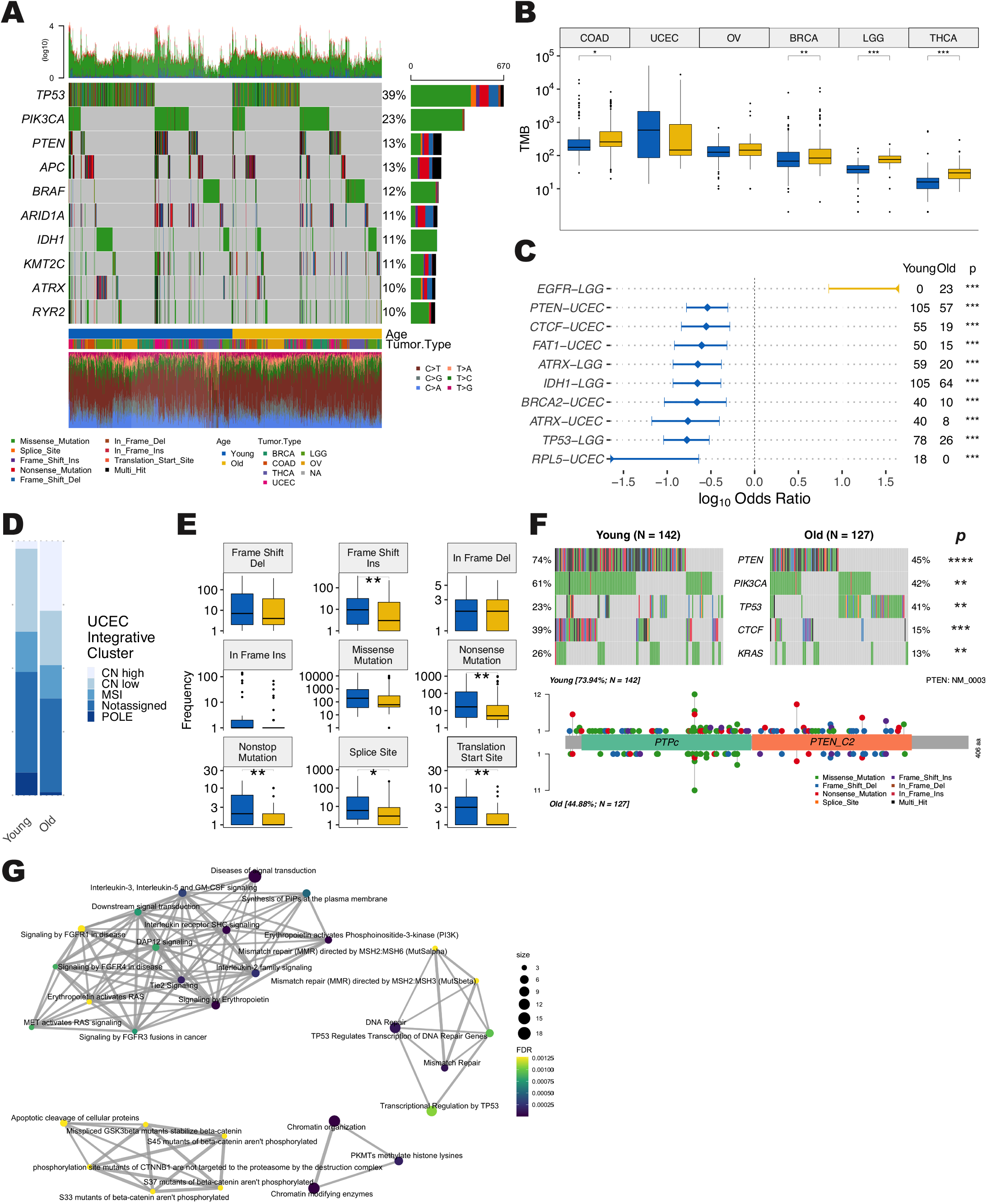
Mutation Analysis. (A) Oncoplot of the top 10 most frequently mutated genes in BRCA, COAD, LGG, OV, THCA, and UCEC. (B) Comparison of tumor mutation burden in younger (blue) and older (yellow) patients. Stars indicate BH-adjusted p values. (C) The forest plot shows tumor-type specific driver gene mutations more commonly found in younger (blue) and older patients (FDR < 0.005). (D) Stacked bar plots show the relative proportion of integrative genomic clusters in younger and older UCEC patients. (E) Boxplots show the frequency of variants in younger (blue) and older (yellow) UCEC patients. Stars indicate BH-adjusted p values. (F) Oncoplot shows mutations that are differentially enriched in younger and older UCEC patients (FDR <0.05) and is accompanied by a lollipop plot of PTEN. (G) Reactome pathway analysis of driver genes mutated in young UCEC patients. Nodes represent the fraction of the number of mutated genes associated with young UCEC patients (FDR < 0.05) and pathway size. Edges represent the number of shared genes across pathways.

Since mutations in multiple driver genes are associated with aggressive clinical features, we hypothesized that tumors from young patients might be enriched for driver mutations, contributing to their aggressive phenotype. In order to test this hypothesis, we stratified tumor type-specific driver mutations (Bailey et al., 2018) by age groups. We found that tumor-type specific nonsynonymous driver gene mutations, with the exception of *EGFR* in LGG, are more common in younger patients from the UCEC and LGG TCGA cohorts (FDR < 0.005, **Fig. 4C**), suggesting once more that younger patients, despite their better outcome, have intrinsically more aggressive tumors, at least in some tumor types. Younger LGG patients were highly enriched for mutations in *TP53, ATRX*, and *IDH1*, and younger UCEC patients were enriched for *PTEN, ATRX, CTCF, BRCA2, RPL5*, and *FAT1* in addition to other genes (Supplementary Table 6). Similarly, younger breast cancer patients in the METABRIC dataset were enriched for TP53 mutations, while older patients were enriched for PIK3CA mutations (**Supplemental Fig. 3A**). However, we could not detect these associations in the TCGA-BRCA dataset. Additionally, younger UCEC patients were enriched for mutations in DNA damage response genes, suggesting homologous recombination defects (**Supplemental Fig. 3B**). Furthermore, we report that older UCEC patients are enriched for the high copy number phenotype from integrative genomic clusters published by Getz et al., 2013 (FDR < 0.0001, **Fig. 4D**).

Next, we stratified UCEC variants by type to better understand functional alterations in younger patients. We found that younger UCEC patients were enriched for frameshift insertions, nonsense, nonstop, splice site, and translation start site mutations (FDR < 0.05, **Fig. 4E**). Interestingly, younger patients were enriched for mutations in the PI3K-PTEN-AKT-mTOR and RTK-RAS signaling pathways, while older patients had more *TP53* mutations (FDR < 0.05, **Fig. 4F**). Enrichment analysis of driver genes more frequently mutated in younger patients revealed four distinct network clusters. These include PIK3CA/RTK-RAS signaling pathways, the beta catenin pathway, DNA-damage response (DDR) pathways, and histone modulatory pathways (FDR < 0.01, **Fig. 4G**, **Supplementary Fig. 3C-F**, Supplementary Table 7). Similar enrichment analysis in old UCEC patients revealed a single cluster of *TP53* associated pathways (FDR < 0.05, **Supplemental Fig. 3G**, Supplementary Table 8), suggesting that tumors from young UCEC patients may have a more heterogeneous mutational landscape.

Taken together, we show the enrichment of genomic aberrations in young patients that results in more aggressive tumors. We find that younger UCEC patients are enriched for mutations, while older patients exhibit a high copy number phenotype. Similarly, younger BRCA patients were associated with *TP53* mutations, while older ones were associated with *PIK3CA* mutations.

## Discussion

Aging is a major risk factor for global morbidity and mortality, particularly for cancer development. While cancers are predominantly diagnosed in older populations aged > 65 years, the increasing frequency of certain tumors in young adults demands immediate attention. With this focus, developing an understanding of aging-related changes in tumors may aid in refining various prevention and treatment options. Recent findings have highlighted the links between aging and tumor biology in specific tumor types(Kim et al., 2020; Osako et al., 2020), however, none have yet performed a multi-omics comparison across tumor types. Here, we performed a systematic analysis of publicly available TCGA Cohorts to elucidate the complex and unique biology of tumors across younger and older age groups. We identified six TCGA tumor types that exhibit an age-associated outcome and molecular phenotype. While the aging-associated effects vary by tumor type, we show that these tumor types exhibit dysregulated molecular aging, which drives several processes involving tumorigenesis and the anti-tumor response. We find that most young patients exhibit accelerated epigenetic aging when compared to healthy counterparts, potentially resulting in impaired cellular function. In addition, we find that younger cancer patients often have a stronger association with aging and senescence-related pathways than older ones.

Further, the survey of downstream biological pathways enriched in tumors as a function of age enables us to uncover potential therapeutic opportunities for younger patients compared to older patients, respectively. The interplay of cellular aging and tumor development in younger patients has complex biology involving genomic and epigenomic defects that govern the interaction of tumor cells with the stroma(Schosserer et al., 2017). Indeed, stabilizing the epigenomic landscape through the use of DNA methyltransferase inhibitors (DNMTi) and histone deacetylase inhibitors (HDACi) serve as powerful anti-tumor tools across tumor types (Christmas et al., 2018; Hull et al., 2016; Rodríguez-Paredes and Esteller, 2011). DNMT and HDAC1/3 inhibition have been shown to be a viable therapeutic strategy for several cancers by inhibiting tumor growth as well as augmenting the anti-tumor immune response. While clinical studies have not assessed differential sensitivity in young and old age groups, our analyses suggest that young patients could exhibit enhanced sensitivity to such treatment regimens. Recent work by Oltra et al. shows that HDAC5 inhibition differentially induces apoptosis in breast cancer cell lines sourced from young patients(Oltra et al., 2020). Epigenomic reprogramming of young breast cancer patients results in age-acceleration at the transcriptional level as well, thereby causing gross functional alterations. We also find that young breast cancer patients are enriched for senescence-associated pathways and that these changes are epigenetically driven. Additionally, we report that young thyroid and endometrial cancer patients are associated with senescence-related pathways at a transcriptional level.

We show that the senescence-associated pathways enriched in younger breast and thyroid cancer patients include SASP, an IL-6 mediated secretory phenotype of persistent senescence involving NF-κB signaling, proteolytic enzymes, growth factors, cytokines, and inflammation, ultimately causing tumor progression, malignant transformation, and proliferation(Di et al., 2014; Gosselin et al., 2009; Krtolica et al., 2001; Malaquin et al., 2013; Mavrogonatou et al., 2020). Additionally, we find that younger breast cancer patients are enriched for oxidative stress-induced senescence, which promotes senescence in SASP fibroblasts, larger tumors, and ultimately SASP(Hiebert et al., 2018). As co-administration of senolytic agents with traditional chemotherapeutic drugs is gaining interest (Fleury et al., 2019; Gayle et al., 2019), our data suggest that tumors from younger patients may be more susceptible to this treatment strategy than older ones.

In addition to a senescent phenotype, we show that younger breast cancer patients are enriched for immune-associated pathways. However, tumors from younger breast cancer patients are often triple negative and are associated with *BRCA1* and *BRCA2* mutations, resulting in aggressive tumors with poor prognosis(Anders et al., 2009; Young et al., 2009) and an increased tumor mutational burden (TMB)(Lal et al., 2019). Several studies have shown a strong link between TMB and the anti-tumor immune response, potentially due to an increased neoantigen burden(Fernandez et al., 2019; Rizvi et al., 2015; Yarchoan et al., 2017). Given the extensive epigenomic aberrations seen in young patients, it is possible that these neoantigens may be epigenetically silenced. This, in turn, would lead to poor prognosis even though the patients are enriched for immune pathways, potentially explaining the increased sensitivity of epigenetic drugs in younger patients(Bell et al., 2018). While the datasets used for this analysis did not have a substantial amount of triple negative cases, future population-based studies could test this hypothesis.

Next, we find that younger endometrial cancer patients are particularly enriched for mutations in several driver genes, including DNA damage response (DDR) genes. Although pathway enrichment analysis of these genes reveals that the PI3K and RTK-RAS pathways are the most mutated, clinical trials targeting these pathways have shown modest success(Dedes et al., 2011; Oda et al., 2005). Mutations in DDR genes of young UCEC patients lead to homologous recombination deficiencies and hypermutator phenotypes. Such populations are sensitive to several therapeutic strategies, including Poly (ADP-Ribose) polymerase 1 inhibition (PARPi)(McCabe et al., 2006). *PTEN* was the most differentially mutated gene in young UCEC patients. PARPi, when coupled with *PTEN* mutations, confers synthetic lethality to such tumors(Mendes-Pereira et al., 2009), suggesting an additional benefit of PARPi in young UCEC patients. Additionally, we show that young LGG patients are enriched for *ATRX* and *IDH1* mutations. Similar to *PTEN* in UCEC, mutations in *ATRX* confer sensitivity to combined PARPi and radiotherapy(Fazal Salom et al., 2018). Furthermore, young LGG patients may be sensitive to *IDH1* inhibitors such as Ivosidenib(DiNardo et al., 2018).

Finally, even though young patients are associated with better survival outcomes, molecular data suggests they are more aggressive tumors that may be restrained by a stronger, more highly activated functional immune system. We found that the gene signatures of immune pathways exhibit higher expression in younger donors, with increased infiltration of B and T cells (the primary cell types associated with anti-tumor immune memory). Therefore, we have begun to characterize the unique biology of tumors in young adults, demonstrate that aging-associated dysfunction is tumor-type specific, and explore the biological systems underlying aggressive tumors. These dysregulated aging and oncogenic processes associated with the aggressive tumors from young patients may be leveraged for differential therapeutic strategies and biomarker discovery.

## Methods

### Age Groups

Previously identified quartiles served as age group limits for individual age-associated tumor types from the TCGA cohort. Additionally, the quartile limits for the TCGA breast cancer cohort was employed to stratify the METABRIC cohort into age groups. The GTEx cohort was, however, classified as young and old using an alternate methodology. Young samples are from individuals younger than 50 years, and old samples are from individuals older than 59 years. This was necessary since age was a discrete variable in the GTEx dataset.

### Differential Gene Expression Analysis

Differentially expressed genes (FDR < 0.05) between young and old age groups were identified across all datasets. In the case of the microarray dataset, *limma* was run with default parameters. For RNA-Seq based data, lowly expressed genes that had less than two counts per million reads in more than two samples were removed from the analysis. The data was voom transformed prior to fitting a linear model using *limma*(Ritchie et al., 2015). Empirical Bayes shrinkage was applied to the model in both cases.

### Pathway Enrichment Analysis

We computed pathway enrichment analysis using GSEA and Reactome pathway analysis. Briefly, GSEA was computed for differential gene expression results using 1000 permutations of hallmark and KEGG pathways using the *fgsea* R package(Sergushichev, 2016). We carried out Reactome pathway enrichment analysis using *clusterProfiler*(Yu et al., 2012). Pathways with an adjusted p-value (Benjamini-Hochberg) less than 0.05 were considered significantly enriched.

### Differential Methylation Analysis

We transformed gene level beta values to m-values using the *wateRmelon* R package(Pidsley et al., 2013) and subsequently identified differentially methylated genes using *limma*.

### ssGSEA Analysis

ssGSEA was estimated using publicly available signatures on MSigDB and the *GSVA* R package(Hänzelmann et al., 2013). The Poisson kernel was employed for microarray and raw count data, while the Gaussian kernel was employed with TPM data. Differential analyses were conducted using the Wilcoxon test and t test.

### DNAm Age Calculation

The epigenetic age for age-dependent cancers, along with normal adjacent tissue (NAT), was imputed using the online DNAm age calculator (https://dnamage.genetics.ucla.edu/home) (Horvath, 2013). This tool predicts age from the DNAm coefficients of 353 CpG sites. Imputations that correlated with the internal gold standard less than 0.8 were discarded, as recommended by the tool, from downstream analyses. Groups were compared using Wilcoxon tests.

### RNA Age Calculation

We used batch corrected FPKM data (TCGA tumor, TCGA NAT and GTEx) obtained from Wang et al. (Wang et al., 2018) to determine transcriptional age using the *RNAAgeCalc* R package(Ren and Kuan, 2020). The regression model for imputing age was the *Dev* signature, which encompassed coefficients for genes that had the largest variation across samples. Groups were compared using Wilcoxon tests.

### Gene Overlap

Pairwise comparison of differentially expressed genes (FDR < 0.05) between young and old samples from each tumor type was performed using Fisher’s exact tests from the *GeneOverlap* R package. P-values were corrected by the Benjamini-Hochberg method. The overlap between hypermethylated (FDR < 0.05) and downregulated genes (FDR < 0.05), and vice versa, was similarly computed.

### Differential mutation analysis

MAF files were obtained from GDC and METABRIC, respectively. We discarded the top 20 frequently mutated genes (FLAGS) in public exomes before proceeding with analyses (Shyr et al., 2014). The enrichment of Single Nucleotide Variants (SNVs) in age groups per tumor type was calculated using Fisher’s exact test from the *maftools* R package(Mayakonda et al., 2018). Tumor mutation burden (TMB) was calculated by dividing the total number of nonsynonymous variants in a sample by genome size (50 MB). SNV classes and TMB were compared across age groups using the Wilcoxon test.

### Data Availability

TCGA clinical data were obtained from Liu et a.l(Liu et al., 2018), CIBERSORT data from Vesteinn et al. (Thorsson et al., 2018), driver gene calls from Bailey et al. (Bailey et al., 2018) and TCGA-UCEC integrative genomic clusters from Getz et al. (Getz et al., 2013). Gene expression data for TCGA samples using RNASeq, methylation profiling data using Illumina 450K, and mutect2 MAF files were downloaded from the Genomic Data Commons. Preprocessed gene-level methylation data were obtained from GDAC. Batch corrected gene expression data were obtained from Wang et al. (Wan g et al., 2018). Clinical data, gene expression by microarray, and MAF files for the Metabric dataset(Curtis et al., 2012; Pereira et al., 2016) were obtained from cbioportal(Gao et al., 2013). Gene expression using RNASeq was obtained from the GTEx consortium v8 release.

### Code Availability

All analyses were performed on R (www.r-project.org) version 3.5.1. Code for all analyses is available on https://github.com/yajass/tcga_aging_final.

**Figure S1:**
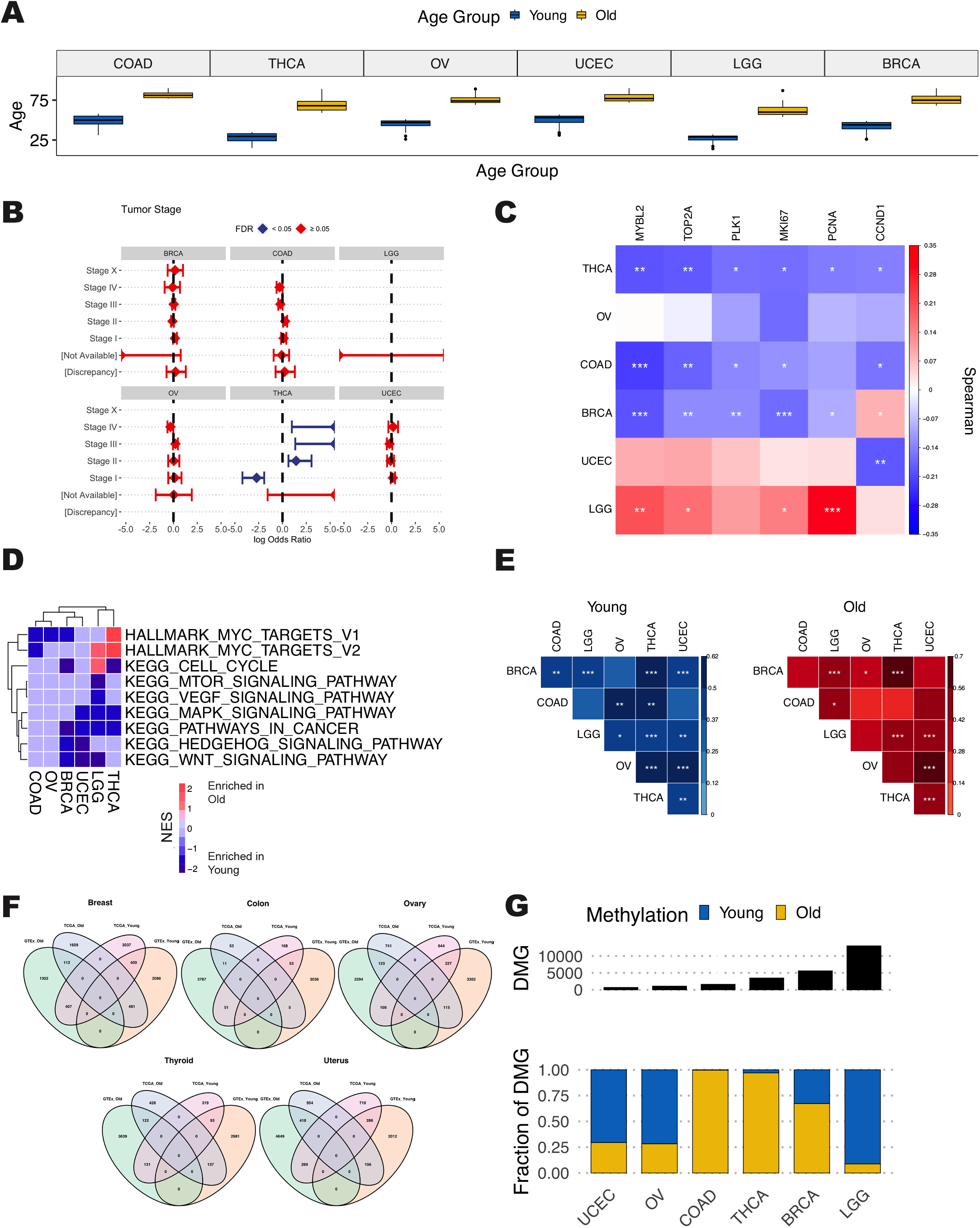
(A) Age distribution that classifies patients as younger and older. (B) The forest plot shows the association between tumor stage and age. Tumor stages with a log odds ratio greater than 0 are associated with older patients. (C) Gene expression correlation coefficients (Spearman) with age for genes associated with tumor progression markers are visualized as a heatmap. Stars indicate BH-adjusted p values. (D) Gene set enrichment analysis of DEG for KEGG pathways associated with tumor proliferation. Pathways in red are associated with older patients, and those in blue are associated with younger patients (FDR < 0.05). (E) The overlap between DEGs associated with young (blue) and old (red) patients visualized as a heatmap. The color gradient reflects log odds ratios, and stars indicate BH-adjusted p values. (F) The Venn diagrams show the overlap between DEGs (FDR < 0.05) associated with young and old individuals across tumor (TCGA) and healthy (GTEx) datasets. (G) The total number of differentially methylated genes (DMG) in younger and older patients are visualized (top). Stacked bar plots represent relative amounts of hypermethylation status in younger (blue) and older (yellow) patients.

**Figure S2:**
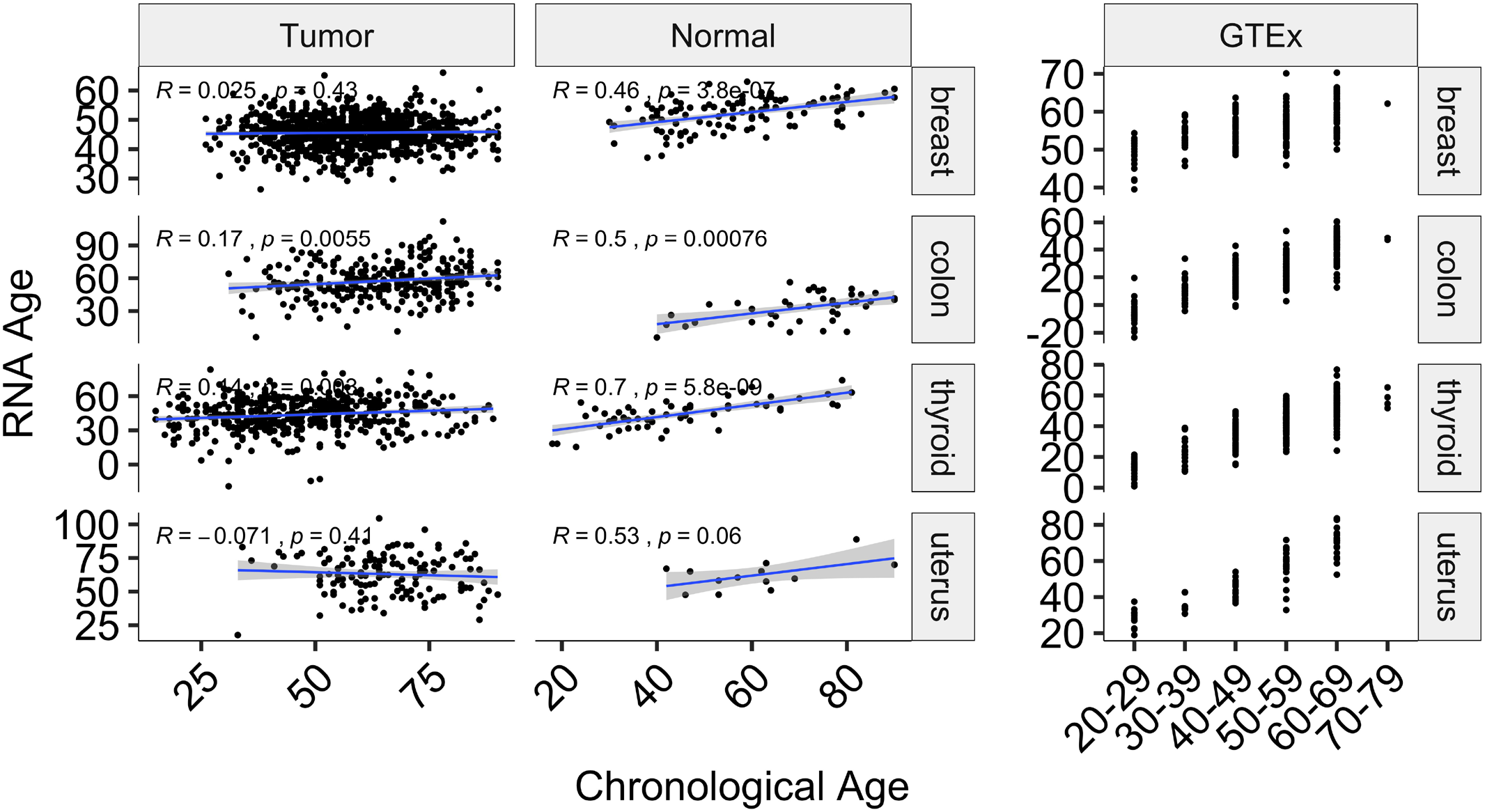
Scatter plots of imputed RNA age and chronological age for TCGA and GTEx datasets

**Figure S3:**
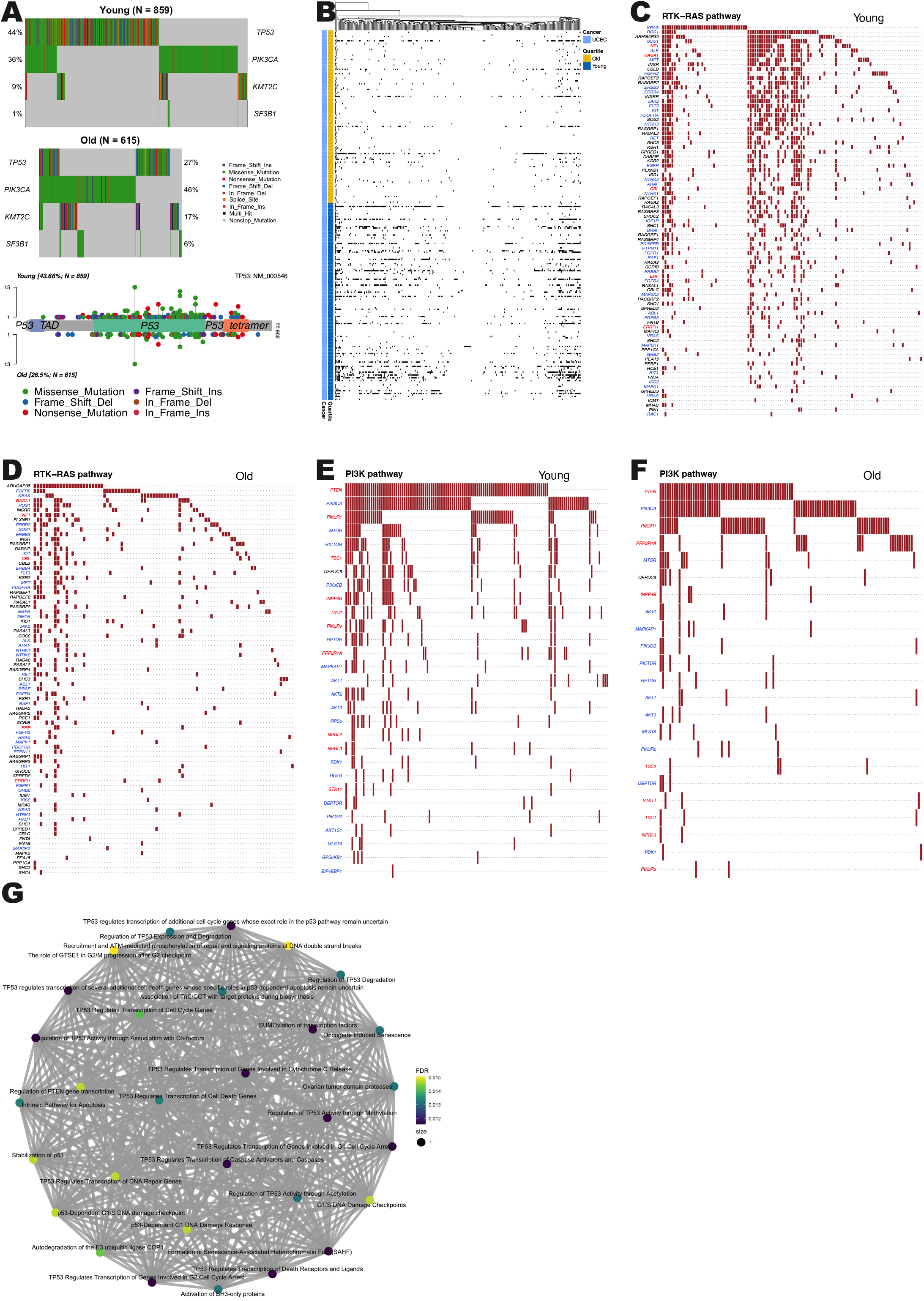
(A) Oncoplot of differential mutations in younger and older patients from the METABRIC dataset (FDR < 0.05) along with a lollipop plot showing the type of P53 variants. (B) The heatmap displays mutation status in DNA damage response genes. Black areas represent mutated genes. (C-F) Oncoplots of the RTK-RAS and PI3K pathways for younger and older patients. Genes in blue are oncogenes, while genes in red are tumor suppressor genes. (G) The network diagram represents reactome pathway enrichment analysis of driver genes in older UCEC patients. Nodes represent the fraction of the number of mutated genes associated with old UCEC patients (FDR < 0.05) and pathway size. Edges represent the number of shared genes across pathways.

## Data Availability

The results in this manuscript are based on publicly available datasets (see Methods).

## Notes

### Competing Interest Statement

The authors have declared no competing interest.

### Funding Statement

No external funding has been received for this work.

### Author Declarations

This research involves deidentified patient data collected by The Cancer Genome Atlas, METABRIC and the Genotype Tissue Expression project. The Weill Cornell Medicine Institutional Review Board found this research exempt since it does not involve identifiable private information.

